# Measuring Tibial Forces is More Useful than Varus-Valgus Laxities for Identifying and Correcting Overstuffing in Kinematically Aligned Total Knee Arthroplasty

**DOI:** 10.1101/19013755

**Authors:** Joshua D. Roth, Stephen M. Howell, Maury L. Hull

## Abstract

Identifying and correcting varus-valgus (V-V) malalignment of the tibial component is important when balancing a kinematically aligned total knee arthroplasty (TKA). Accordingly, the primary objective was to determine whether the tibial forces or V-V laxities are more sensitive to, and thus more useful for identifying and correcting, V-V malalignments of the tibial component that overstuff a compartment. Calipered kinematically aligned TKA was performed on nine human cadaveric knees. Medial and lateral tibial forces and V-V laxities were measured from 0° to 120° flexion with an unmodified reference tibial component and modified tibial components that introduced ±1° and ±2° V-V malalignments from the reference component to overstuff either the medial or lateral compartment. Changes in the tibial forces were most sensitive to V-V malalignments at 0° flexion (medial = 118±34 N/deg valgus malalignment and lateral = 79±20 N/deg varus malalignment). The varus and valgus laxities were most sensitive to V-V malalignments at 30° flexion (−0.6±0.1 deg/deg varus malalignment) and 120° flexion (−0.4±0.2 deg/deg valgus malalignment), respectively. The maximum average signal-to-noise ratios of the sensitivities to changes in tibial forces and V-V laxities were 8.4 deg^-1^ and 0.9 deg^-1^, respectively, based on reported measurement errors (i.e., noise) using current intraoperative technologies (14 N and 0.7°). Because of the greater signal-to-noise ratios, measuring tibial forces is more useful than V-V laxities for identifying and correcting V-V malalignments of the tibial component that overstuff a compartment.

**Clinical Significance:** The sensitivities of tibial forces provide objective guidance to surgeons performing V-V recuts of the tibia.

## Introduction

The surgical alignment goal of kinematically aligned total knee arthroplasty (TKA) is to restore the native joint lines with the ultimate functional goal of restoring knee function to native. Restoration of the joint lines, in turn, closely restores the native alignments of the limb and knee and eliminates the need for ligament releases^1, 2^. Kinematically aligning the femoral component is relatively straight forward because the resection thicknesses can be measured using calipers (i.e., calipered kinematic alignment^3^). However, kinematically aligning the tibial component is more challenging because the resection plane must be correctly aligned in both the coronal and sagittal planes. The combination of irregular geometry of the tibial articular surface^4^ and large amounts of cartilage and bone wear^5^ further complicates the task of achieving correct alignment of the tibial resection plane.

In calipered kinematically aligned TKA, the varus-valgus (V-V) orientation of the tibial resection is adjusted to balance medial and lateral soft tissues. The current intraoperative verification check for proper V-V alignment of the tibial component is that the knee is stable in extension based on a manual V-V laxity assessment by the surgeon^6^. However, because identifying an overly tight (i.e., overstuffed) knee by manually checking laxities is challenging^7^, it is important to consider more accurate intraoperative technologies such as computer navigation for measuring laxities^8^ and sensors for measuring tibial forces^9, 10^. The utility of these two technologies can be compared based on their signal-to-noise ratios. The ‘signal’ is the sensitivity of changes in the laxities/tibial forces due to V-V malalignment of the tibial component that overstuffs a compartment of the tibiofemoral joint. The ‘noise’ is the error in measuring the laxities/tibial forces using current intraoperative technologies. Thus, the better of the two technologies for detecting V-V malalignments of the tibial component would have a higher signal-to-noise ratio. Further, the sensitivities of changes in the better variable would indicate to the surgeon the amount of adjustment necessary in the V-V orientation of the tibial resection to realize a desired change in laxity or change in tibial force to achieve the desired soft tissue balance.

Prior studies have indicated that changes in the tibial forces are sensitive to small V-V malalignments of the tibial component in mechanically aligned TKA^11^. However, because there are fundamental differences between the surgical alignment goals and surgical techniques to achieve the surgical alignment goals between mechanically and kinematically aligned TKA^12^, it is important to determine the sensitivity in changes in both the laxities and tibial forces to V-V malalignments of the tibial component in kinematically aligned TKA. Additionally, because prior studies have shown wide variability in both the tibial forces^13^ and laxities^14^ from knee to knee, it is important to determine how much these sensitivities vary from knee to knee. If the sensitivities vary widely, then it might not be feasible to determine the amount of correction for an individual patient based on a mean value.

Accordingly, there were three objectives. The first objective was to determine the sensitivities of the tibial forces and V-V laxities to V-V malalignments of the tibial component that overstuff a compartment. By comparing signal-to-noise ratios for V-V laxities and tibial forces based on sensitivities and errors using currently available technologies^15, 16^, the second objective was to determine whether the changes in laxities or tibial forces are more useful for identifying and correcting V-V malalignments of the tibial component that overstuff a compartment. The third objective was to determine whether the variability in sensitivities from knee to knee was sufficiently low so that the amount of correction for an individual could be based on the mean value of the sensitivity.

## Methods

Nine fresh-frozen human cadaveric knees (average age = 74 years, range = 57 to 93 years, 6 males) were included. With nine knees, strong relationships between V-V malalignments of the tibial component and tibial forces/laxities (i.e., r^2^ > 0.55) could be detected with α = 0.05 and (1-β) = 0.8^17^. Each knee was free of radiographic signs of degenerative joint disease (i.e., marginal osteophytes, joint space narrowing, chondrocalcinosis, and/or subchondral sclerosis) and evidence of previous surgery to the knee.

After thawing overnight, each native knee specimen was dissected and aligned in a six degree-of-freedom load application system^18^ in preparation for measuring the tibial forces and laxities using previously described protocols^14, 19-22^. Briefly, to enable the application of muscle forces, cloth loops were sutured to the tendons of insertion of the quadriceps, the biceps femoris, and the semimembranosus and semitendinosus (sutured together due to shared line of action^23^). Subsequent to a functional alignment procedure^14, 19^, the shafts of the femur and tibia were cemented within square aluminum tubes using methyl methacrylate (COE Tray Plastic, GC America Inc., Alsip, IL) to rigidly fix the position and orientation of the knee specimen relative to the load application system and enable removal and reinsertion of the knee specimen during subsequent testing in the load application system^18^. Before determining the full extension position of each knee (i.e., 0° flexion), each knee specimen was subjected to a preconditioning protocol consisting of first cycling five times between ± 2.5 Nm in flexion–extension (F–E) and then extending under 2.5 Nm to define 0° flexion^24^.

Kinematically aligned TKA was performed using cruciate-retaining components (Persona CR, Zimmer Biomet, Inc, Warsaw, IN, USA) and disposable manual instruments without ligament release following a previously described technique^3, 25^. Briefly, the knee was exposed through a mid-sagittal osteotomy of the patella^26^. The goal of maintaining the native the distal and posterior femoral joint lines was accomplished by matching the thicknesses of the distal medial, distal lateral, posterior medial, and posterior lateral femoral resections to within 0.5 mm as measured using calipers to the corresponding condylar regions of the femoral component after correcting for the kerf of the saw blade^25^.

Next a conservative tibial resection was performed with the V-V orientation and posterior slope set visually by the surgeon to match the those of the native tibial plateau. After inserting trial components, the V-V orientation of the tibial resection was adjusted until there was negligible V–V laxity at 0° flexion as judged visually by the surgeon^19, 25^. The posterior slope of the tibial resection was also adjusted via re-cuts until the anterior-posterior (A–P) offset (i.e., distance between the distal medial femoral condyle and the anterior cortex of the tibia) measured with a caliper at 90° flexion matched that of the knee at the time of exposure^25^. These checks have been shown to result in a V-V orientation of the tibial component within 0.0° ± 1.8° of that of the contralateral healthy knee^2^ and a posterior slope within −0.2° ± 2.5° of the pre-operative posterior slope^1^. The internal-external (I–E) rotation of the tibial component was set parallel to the F–E plane of the knee, which is the plane in which the tibia flexes and extends, using templates that have been shown to align the A–P axis of the tibial component with a root mean squared error of 4° from the F–E plane of the knee^27^.

After the correctly sized trial components were determined and properly aligned based on the previously described verification checks, the femoral component was coated with petroleum jelly and cemented to the distal femur. The tibial baseplate was coated with petroleum jelly and cemented into the proximal tibia after which the correctly sized tibial insert was attached. The petroleum jelly allowed the components to be released from the cement mantles, which enabled accurate exchanges of tibial components and the tibial force sensor described below. After the cement cured, the components were removed. The patella was not resurfaced.

Varus–valgus malalignment of the tibial component was created by modifying both a commercially available tibial baseplate (Persona CR, Zimmer Biomet, Inc.) for use while measuring the laxities and the articular surface inserts of the custom tibial force sensor^21^ for use while measuring the tibial forces (Figure 1). These modifications were performed using 3D modeling software (SolidWorks 2014, Dassault Systèmes). To create malalignments that would be correctable by a V-V re-cut, varus malalignments were made about the medial edge of the tibial baseplate, and valgus malalignments were made about the lateral edge of the tibial baseplate. Thus, varus malalignments primarily overstuffed the lateral compartment, and valgus malalignments primarily overstuffed the medial compartment (Figure 1). Five tibial baseplates and five sets of articular surface inserts were 3D printed (Gray60, Objet Eden260VS, Stratasys, Ltd.) for each tibial baseplate size (four with modified geometry to create malalignments of 1° varus, 2° varus, 1° valgus, and 2° valgus and one 0° reference baseplate/set of articular surface inserts with unmodified geometry). Values of 1° and 2° were selected to span the malalignments correctable with 2° re-cut guides.

**Figure 1.**
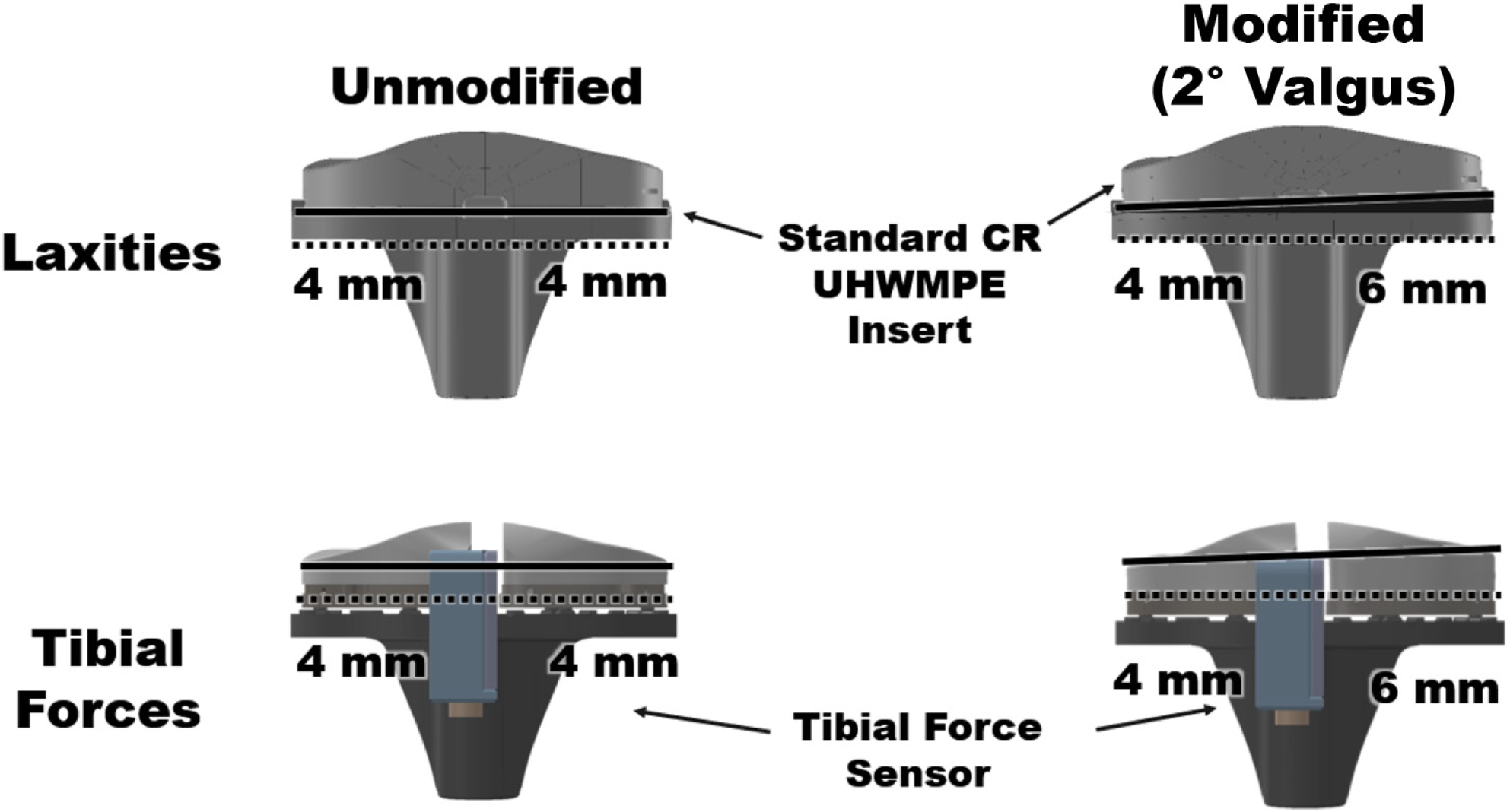
Renderings show the design of the modified tibial baseplates used when measuring the laxities (top row) and the design of the modified articular surface inserts used when measuring the tibial forces (bottom row). For the tibial baseplate, the modifications consisted of rotating the proximal features of the baseplate that secured the insert in place (solid black line) relative to the distal surface of the baseplate that interfaced with the cement mantle (dotted black line). Because the geometry of the distal surface was not changed, all baseplates could be implanted using the same cement mantle in each knee. The geometry of the proximal locking features of the tibial baseplate was also unchanged so the same standard UHMWPE (ultra-high molecular weight polyethylene) liner could be used in all knees. For the articular surface inserts, the modifications consisted of rotating the proximal articular surface (solid black line) relative to the distal surface that secured the inserts to the tibial force sensor (dotted black line)^*21*^. The modified tibial baseplate and articular surface inserts (left column) are an example of one of the four modified tibial baseplate and articular surface inserts created for each size tibial component. In the example case of a 2° valgus malalignment, the medial side of the tibial component is approximately 2 mm thicker than the lateral side.

Tibial forces in the medial and lateral compartments were measured with each of the five sets of 3D-printed articular surface inserts attached to the custom tibial force sensor^21^. The tibial force sensor had the same exterior size and shape as the correctly sized Persona tibial baseplate and insert. The tibial force sensor measured force independently in the medial and lateral compartments and over the full surface area of the liner with a maximum root mean squared error of 6 N^21; 39^. Before testing, each knee was passively flexed five times between 0° and 120° of flexion. The testing order of the five sets of 3D-printed articular surface inserts was randomized. After inserting a randomly selected set of 3D-printed articular surface inserts, the patellar osteotomy was closed with two transverse screws. To stabilize the TKA knee during flexion, constant forces of 26 N, 80 N, and 15 N were applied to the tendons of insertion of the semimembranosus/semitendinosus, quadriceps, and biceps femoris, respectively. These forces were proportional to the muscle cross-sectional areas^28^ and smaller than forces used to stabilize the TKA in other studies^29-36^. The tibial forces in the medial and lateral compartments were measured at 30° increments as the TKA was flexed passively from 0° to 120° and back to 0°. After a test was completed for a set of 3D-printed articular surface inserts, the patellar osteotomy was opened, a different set of 3D-printed articular surface inserts was inserted, the patellar osteotomy was closed, and the test was repeated.

Next the varus and valgus laxities were measured with each of the five 3D-printed tibial baseplates using a six degree-of-freedom load application system^14, 18, 22^. With the correctly sized components implanted, each knee was subjected to a preconditioning protocol consisting of first passively flexing and extending the knee five times from 0° to 120° flexion. Next, the knee was moved to a flexion angle randomly selected from 0°, 60°, and 120° and then cycled five times between the prescribed load limits for in four degrees of freedom in a random order^37^. The prescribed load limits were ± 3 Nm for I–E rotation^38^, ± 5 Nm for V–V rotation^39^, ± 45 N for A–P translation^40^, and ± 100 N for compression-distraction translation^41^. The limits of each load were selected to engage the soft tissues sufficiently to load them beyond the initial toe region of the tibiofemoral joint’s load– deformation curve^39, 40^. The protocol was repeated for the remaining two flexion angles. After completing preconditioning, the V-V laxities were measured at 30° increments from 0° to 120° in a random order. The V-V laxities were computed as the difference between the V-V position of the tibia under ± 5 Nm and that in the unloaded knee^22^.

Repeatability of laxities and tibial forces was determined by measuring each variable over five trials in three additional knee specimens. The greatest standard deviations of the V-V laxities and tibial forces were 0.1°^22^ and 12.9 N^20^, respectively.

Following University of California policies, this study did not require institutional review board (IRB) approval because de-identified cadaveric specimens were used.

### Statistical analysis

To determine the sensitivities of the tibial forces and V-V laxities to V–V malalignment of the tibial component, a simple linear regression with the intercept set to zero was performed for the changes in the tibial forces and V-V laxities at each flexion angle. The changes were computed as the difference between the tibial force/laxity with each modified component and that with the unmodified reference component. The degree of either varus or valgus malalignment of the tibial component was the independent variable, and the changes in each tibial force/laxity in all nine knees were the dependent variables. The regressions were performed separately for varus malalignments and valgus malalignments because each is likely to affect the medial and lateral structures differently due to the differences in stiffness of the soft tissue restraints^36, 38, 42^. The slope of each regression line determined the sensitivity of each tibial force variable/laxity to each direction of V-V malalignment of the tibial component at that flexion angle.

To compare the sensitivities of tibial forces and V-V laxities to V–V malalignments of the tibial component, the signal-to-noise ratios (i.e., absolute values of the ratios of the sensitivities to previously reported measurement errors of either optical motion capture systems or intraoperative tibial force sensors) were computed. Measurement errors (i.e., noise levels) of 0.7° were used for the laxities^15^. Measurement errors (i.e., noise level) of 14 N^16^ were used for the tibial forces. The signal-to-noise ratios were computed for each sensitivity at each flexion angle.

To determine the variability of the sensitivities of the tibial forces and V-V laxities to V–V malalignment of the tibial component, the 95% tolerance limits, which bound the predicted range of 95% of the sensitivities in the population with a 95% confidence level^43^, were computed for the sensitivity of each variable at each flexion angle (Equation 1). With nine knee specimens included in the present study, the tolerance factor (*k*) was computed to be 3.55^43^. Also, a simple linear regression was performed for each knee independently with the degree of either varus or valgus malalignment of the tibial component as the independent variable and the changes in each tibial force/laxity in each knee as the dependent variable.

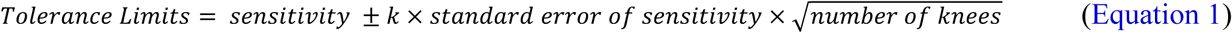

## Results

Sensitivities of tibial forces to V-V malalignment of the tibial component were greatest at 0° flexion (Table 1, Figure 2). Valgus malalignments caused changes in the tibial forces throughout flexion, but varus malalignments caused changes only at 0° flexion. Valgus malalignments primarily increased the medial tibial force. Varus malalignments primarily increased the lateral tibial force at 0° flexion.

**Table 1.** Mean sensitivity (i.e. slope), coefficient of determination (r^2^), p-value, and 95% tolerance limits of the sensitivity for linear regressions between the change in the medial and lateral tibial forces and either the degree of varus or valgus malalignment of the tibial component (TC) (independent variables).

|  |  | 0° flexion | 30° flexion | 60° flexion | 90° flexion | 120° flexion |
| --- | --- | --- | --- | --- | --- | --- |
| Sensitivity of Lateral Tibial Force to Varus Malalignment | Slope | 78.6 N/deg | 7.2 N/deg | 4.9 N/deg | 5.5 N/deg | 4.4 N/deg |
| | $r^2$ | 0.68 | 0.39 | 0.52 | 0.48 | 0.43 |
|  | p-value | <0.0005 | <0.0005 | <0.0005 | <0.0005 | <0.0005 |
|  | 95% tolerance limits | 7.7 to 149.6 N/deg | -4.7 to 19.1 N/deg | -1.4 to 11.2 N/deg | -2.1 to 13.1 N/deg | -2.2 to 11.0 N/deg |
| Sensitivity of Medial Tibial Force to Valgus Malalignment | Slope | 118.3 N/deg | 35.9 N/deg | 39.5 N/deg | 30.4 N/deg | 18.2 N/deg |
| | $r^2$ | 0.63 | 0.49 | 0.53 | 0.44 | 0.24 |
|  | p-value | <0.0005 | <0.0005 | <0.0005 | <0.0005 | <0.0005 |
|  | 95% tolerance limits | -1.4 to 238.0 N/deg | -12.9 to 84.7 N/deg | -10.1 to 89.1 N/deg | -15.0 to 75.7 N/deg | -25.0 to 61.5 N/deg |

**Figure 2.**
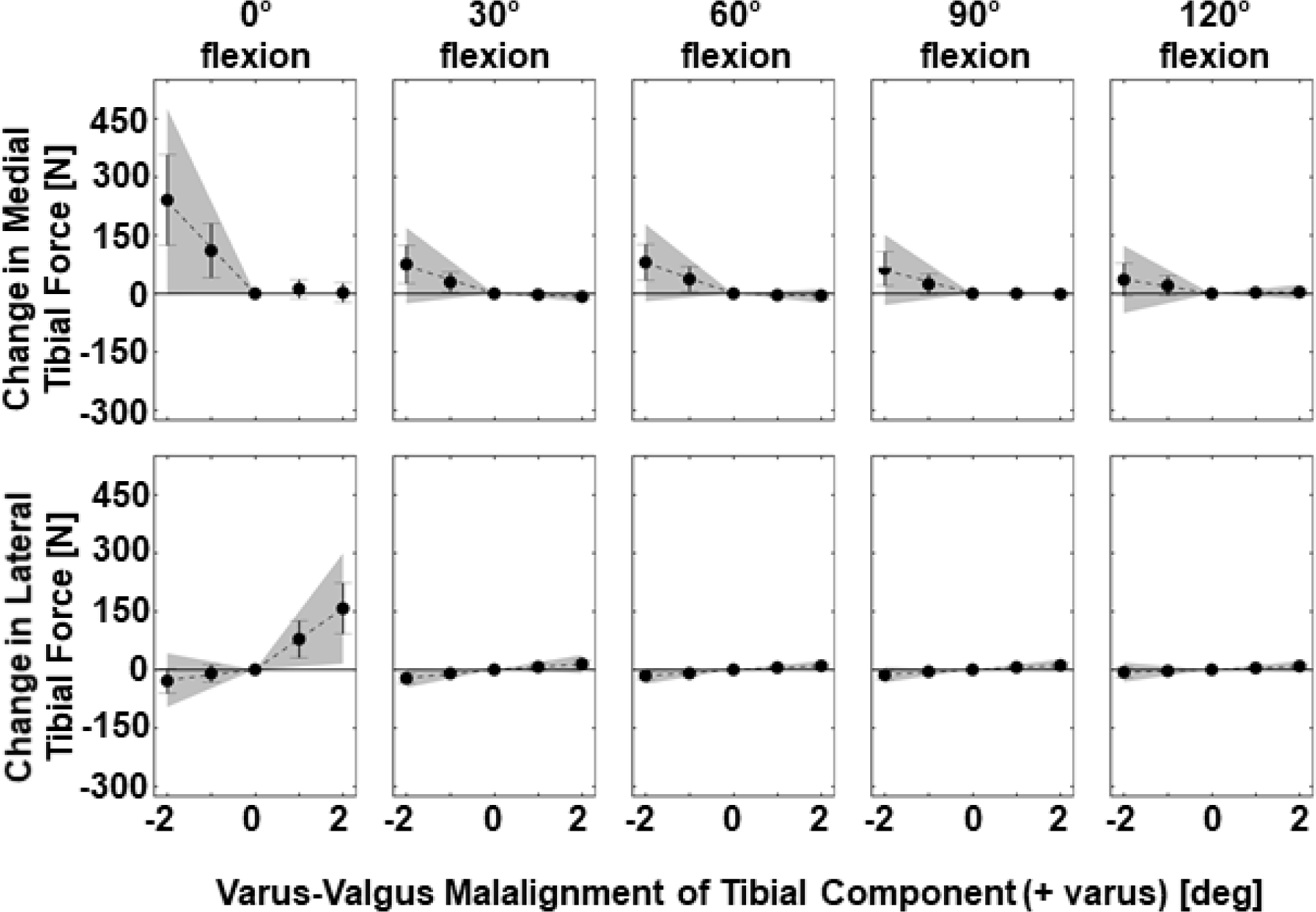
Plots show means (points) and standard deviations (error bars) of the changes in the medial and lateral tibial forces caused by V-V malalignment of the tibial component. Dotted lines show the significant relationships (p < 0.05) between the changes in tibial force variables and V-V malalignments (Table 1). The shaded grey regions show the 95% tolerance intervals for the sensitivities.

Unlike the sensitivities of tibial forces, which were greatest at 0° flexion, the sensitivities of V-V laxities to V-V malalignment were greatest in mid to deep flexion (Table 2). Valgus malalignments decreased both varus and valgus laxities with the greatest sensitivity occurring at 120° flexion for valgus laxity. Varus malalignments primarily decreased the varus laxities with the greatest sensitivity occurring at 60° flexion.

**Table 2.** Mean sensitivity (i.e. slope), coefficient of determination (r^2^), p-value, and 95% tolerance limits of the sensitivity for linear regressions between the changes in the laxities (dependent variables) and the degree of either varus or valgus malalignment of the tibial component (TC) (independent variables).

|  |  | 0° flexion | 30° flexion | 60° flexion | 90° flexion | 120° flexion |
| --- | --- | --- | --- | --- | --- | --- |
| Sensitivity of Varus Laxity to Varus Malalignment | Slope | -0.2 deg/deg | -0.6 deg/deg | -0.6 deg/deg | -0.6 deg/deg | -0.6 deg/deg |
| | $r^2$ | 0.39 | 0.88 | 0.80 | 0.68 | 0.63 |
|  | p-value | <0.0005 | <0.0005 | <0.0005 | <0.0005 | <0.0005 |
|  | 95% tolerance limits | -0.7 to 0.2 deg/deg | -0.9 to -0.3 deg/deg | -1.0 to -0.2 deg/deg | -1.2 to -0.1 deg/deg | -1.2 to 0.0 deg/deg |
| Sensitivity of Valgus Laxity to Valgus Malalignment | Slope | -0.1 deg/deg | -0.3 deg/deg | -0.1 deg/deg | -0.2 deg/deg | -0.4 deg/deg |
| | $r^2$ | 0.20 | 0.30 | 0.05 | 0.24 | 0.44 |
|  | p-value | <0.0005 | <0.0005 | 0.09 | <0.0005 | <0.0005 |
|  | 95% tolerance limits | -0.5 to 0.2 deg/deg | -0.8 to 0.3 deg/deg | -0.9 to 0.6 deg/deg | -0.7 to 0.3 deg/deg | -1.1 to 0.2 deg/deg |

The signal-to-noise ratios of the tibial forces were often greater than those of the V-V laxities (Figure 3). For the tibial forces, the greatest ratios occurred at 0° flexion. These were 5.6 deg^-1^ of varus malalignment for the lateral tibial force and 8.5 deg^-1^ of valgus malalignment for the medial tibial force. In comparison, for the V-V laxities, the greatest ratios were 0.9 deg^-1^ of varus malalignment for the varus laxity at 90° of flexion and 0.6 deg^-1^ of valgus malalignment for the valgus laxity at 120° of flexion (Figure 3).

**Figure 3.**
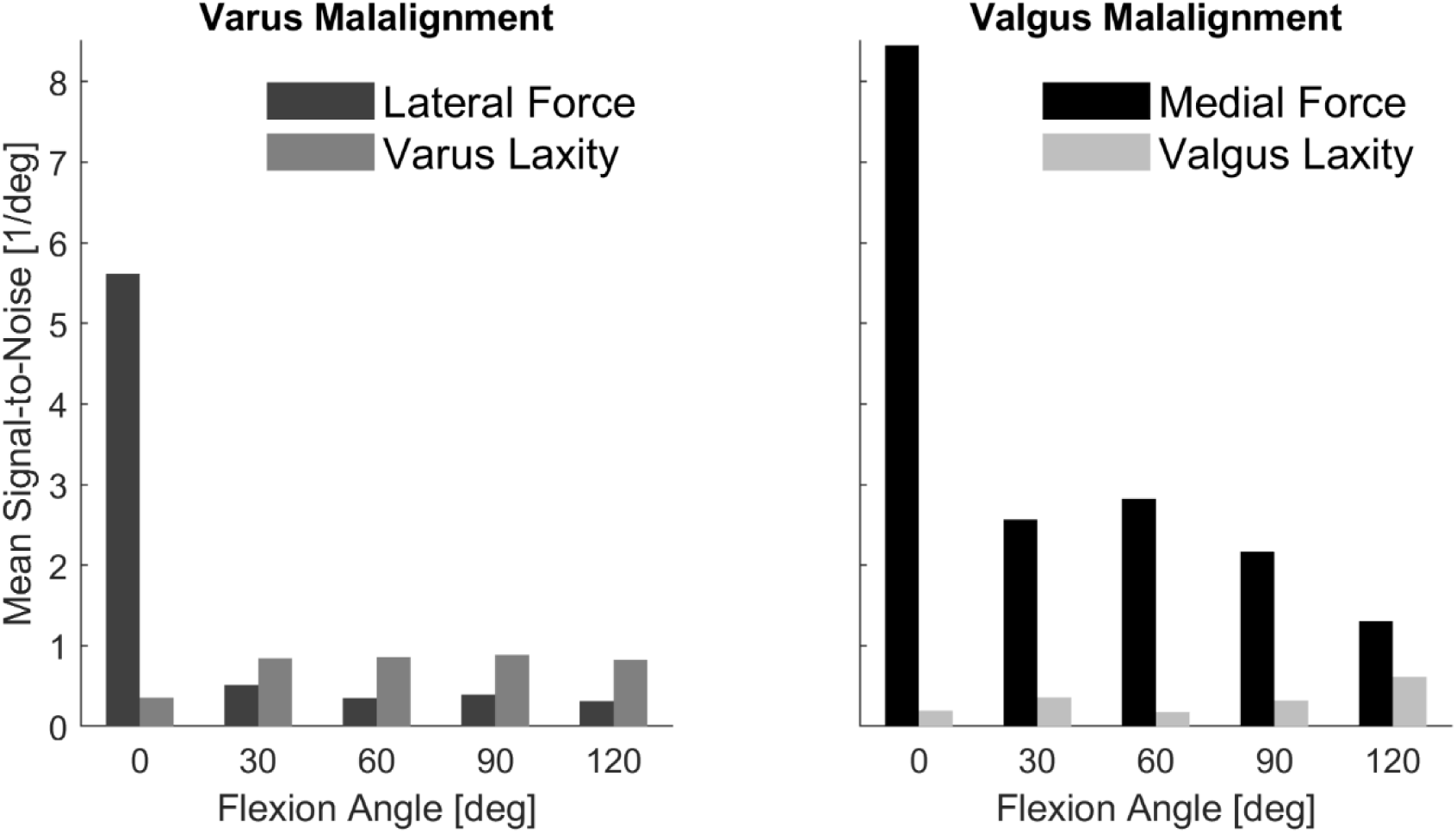
Bar plots show the mean signal-to-noise ratios (i.e., ratios of the sensitivities to previously reported measurement errors of either optical motion capture systems or intraoperative tibial force sensors) for the medial and lateral forces, and the varus and valgus laxities from 0° to 120°.

The sensitivities of both tibial forces and laxities to V-V malalignment varied widely from knee to knee (Tables 1 and 2). For the tibial forces, the 95% tolerance limits of the sensitivities were widest at 0° flexion (medial tibial force = −1 to 238 N/deg of valgus malalignment; lateral tibial force = 8 to 150 N/deg of varus malalignment) (Table 1, Figure 2). For the varus laxities, the 95% tolerance limits for the sensitivities were widest at 90° flexion (−1.2 to −0.1 deg/deg of varus malalignment) (Table 2). For the valgus laxities, the 95% tolerance limits for the sensitivities were widest at 120° flexion (−1.1 to 0.2 deg/deg of valgus malalignment) (Table 2).

Although the sensitivities varied widely from knee to knee, the strength of the linear relationships did not. The coefficients of determination (r^2^) for the sensitivities of tibial forces across the nine knees were most consistent at 0° flexion (range medial tibial force to valgus malalignment = 0.80 to 1.00 and range lateral tibial force to varus malalignment = 0.83 to 1.00) (Table 3). The r^2^-values for the sensitivities were most consistent across the nine knees for the V-V laxities at 60° flexion (range varus laxity to varus malalignment = 0.97 to 1.00, range valgus laxity to valgus malalignment = 0.58 to 1.00).

**Table 3.** Mean sensitivities and coefficients of determination (r^2^) for linear regressions between the change in the medial and lateral tibial forces (dependent variables) and either the degree of varus or valgus malalignment of the tibial component (TC) (independent variables) for each of the nine knees at 0° flexion.

| Specimen | Mean slope, ($r^2$) | | | |
| --- | --- | --- | --- | --- |
|  | Medial Tibial Force<br>[N/deg] |  | Lateral Tibial Force<br>[N/deg] |  |
|  | TC<br>Varus | TC<br>Valgus | TC<br>Varus | TC<br>Valgus |
| 1 | 21.5<br>(0.65) | -212.9<br>(1.00) | 63.9<br>(1.00) | 3.8<br>(0.23) |
| 2 | -13.4<br>(0.91) | -87.9<br>(0.80) | 116.0<br>(0.97) | 26.6<br>(0.86) |
| 3 | 5.5<br>(0.97) | -86.3<br>(1.00) | 52.6<br>(0.97) | 15.2<br>(0.93) |
| 4 | 4.6<br>(0.08) | -179.6<br>(0.99) | 145.4<br>(0.95) | 17.8<br>(0.73) |
| 5 | 13.9<br>(0.96) | -142.1<br>(1.00) | 60.5<br>(0.97) | 17.1<br>(0.16) |
| 6 | 20.1<br>(0.79) | -147.8<br>(1.00) | 30.8<br>(0.83) | 24.3<br>(1.00) |
| 7 | -12.9<br>(0.84) | -35.3<br>(0.99) | 67.1<br>(1.00) | 0.3<br>(0.06) |
| 8 | -13.1<br>(1.00) | -128.4<br>(1.00) | 86.0<br>(1.00) | 24.8<br>(1.00) |
| 9 | 3.2<br>(0.93) | -44.5<br>(0.94) | 85.2<br>(0.99) | -8.9<br>(0.19) |

## Discussion

There were three key findings in the present study. The first key finding was that in general the tibial forces were most sensitive to overstuffing near extension, and the laxities were most sensitive in mid to deep flexion. The second key finding was that tibial forces are better than laxities for identifying and correcting V-V malalignments of the tibial component that overstuff a compartment because the signal-to-noise ratios are greater by about an order of magnitude. The third key finding was that there was wide variability in the sensitivities of both the tibial forces and the laxities to V-V malalignments between knees; however, each knee demonstrated strong linear relationships between changes in tibial force/laxities and V-V malalignments.

The first key finding that tibial forces were most sensitive to overstuffing near extension and the laxities were most sensitive in mid to deep flexion is consistent with the expected changes in the tension of the soft tissue restraints. Overstuffing increases the length of the soft tissue restraints in the overstuffed compartment, which in turn can increase the soft tissue tension or decrease the laxity depending on the initial V-V laxity. If the initial V-V laxity is minimal, thus indicating some degree of pretension in the soft tissue restraints, then overstuffing will further increase the tension and thus the tibial force, which was observed at 0° flexion where V-V laxity is minimal^22^; the V-V laxities will be relatively unaffected because the soft tissue restraints are stiff. However, as the V-V laxity increases with flexion^22^ indicating that the soft tissue restraints become more lax, the effect of overstuffing on tibial forces is mitigated, but the effect on V-V laxities is amplified because overstuffing makes the knee less lax in the particular compartment overstuffed.

Regarding the second key finding, the signal-to-noise ratio indicates how well actual changes in force/laxity can be detected in the presence of measurement errors. Because all the signal-to-noise ratios for the changes in the V-V laxities were less than 1 (Figure 3), surgeons would have a difficult time detecting actual changes from measurement errors. However, many of the signal-to-noise ratios for changes in tibial forces, especially those at 0° flexion where the tibial forces w greatest^20^, are much greater than 1. The relatively large signal-to-noise ratios indicate that actual changes in tibial forces due to V-V malalignments of the tibial component should dominate over measurement errors (Figure 3). Therefore, tibial forces are better than V-V laxities for identifying and correcting V-V malalignments of the tibial component that overstuff a compartment because guiding re-cuts based on the sensitivities in tibial forces should lead to more accurate re-cuts.

The second key finding that the tibial forces are more useful than laxities for identifying and correcting V-V malalignments of the tibial component that overstuff a compartment is supported by prior clinical and biomechanical studies. A previous clinical study showed that it is challenging for surgeons to detect high tibial forces using a standard gap-balancing approach for mechanically aligned TKA^7^, which is based on the joint laxity. Without the use of an intraoperative tibial force sensor, TKAs deemed to be correctly balanced exhibited mean compartment loads as high as 352 N, which are much greater than those in the native knee that have been reported as 114 N on average^13^. Biomechanically, because the soft tissues restraints are often already loaded into their linear high stiffness regions near full extension^39, 40^, additional deformation due to overstuffing does not change their stiffness. Thus, the laxity, which is a cumulative measure of the deformations of many soft tissue restraints, would also not change (Table 2). Therefore, intraoperative laxity assessments are likely better for detecting an under-stuffed compartment where gaps between the components can be detected visually.

Regarding the third key finding, the large but variable sensitivities of tibial forces to overstuffing caused by V-V malalignments of the tibial component have important clinical implications for balancing a kinematically aligned TKA. The wide tolerance intervals on the sensitivities are supported by both the wide variability in the mechanical properties of the primary soft tissue restraints^44-46^ and the wide variability in the tibial forces in the native knee^13^. As described previously, balancing of the medial and lateral soft tissues in calipered kinematically aligned TKA is performed by adjusting the V-V orientation of the tibial resection after confirming that the femoral component is kinematically aligned^19, 25^. Thus, the sensitivities determined in the present study provide quantitative guidance for how large of an adjustment is necessary to achieve a desired reduction in the tibial forces when an intraoperative tibial force sensor is used. However, the wide variability of these sensitivities raises concern with using the mean values for all knees. An illustrative example about how this variability might increase the time and complexity of the balancing procedure is presented in Supplement 1.

Because there was a strong linear relationship between changes in tibial forces and V-V malalignments in all nine knees, a simple procedure might be useful to determine a patient-specific sensitivity (Figure 4) to overcome the wide patient-to-patient variability. In this procedure, the surgeon would first record the initial tibial force values. For example, assume that the initial medial and lateral tibial forces are 300 and 65 N, respectively, and the surgeon desires to match the mean tibial forces of the native knee (114 N in the medial compartment and 63 N in the lateral compartment^13^) by performing a varus re-cut. Next, the surgeon would insert a trial that simulates a 1° varus re-cut and again record the medial and lateral tibial forces. The difference between the medial tibial force before and after inserting the 1° varus re-cut trial is the sensitivity of the medial tibial force to valgus tibial malalignment. By determining this patient-specific sensitivity, the re-cut also would be patient-specific (Figure 4). This approach would be useful in patients with OA because it is based on the current state of the joint; thus, utilizing this simple patient-specific approach would overcome one methodological issue with this study described below which is that the knees did not have OA.

**Figure 4.**
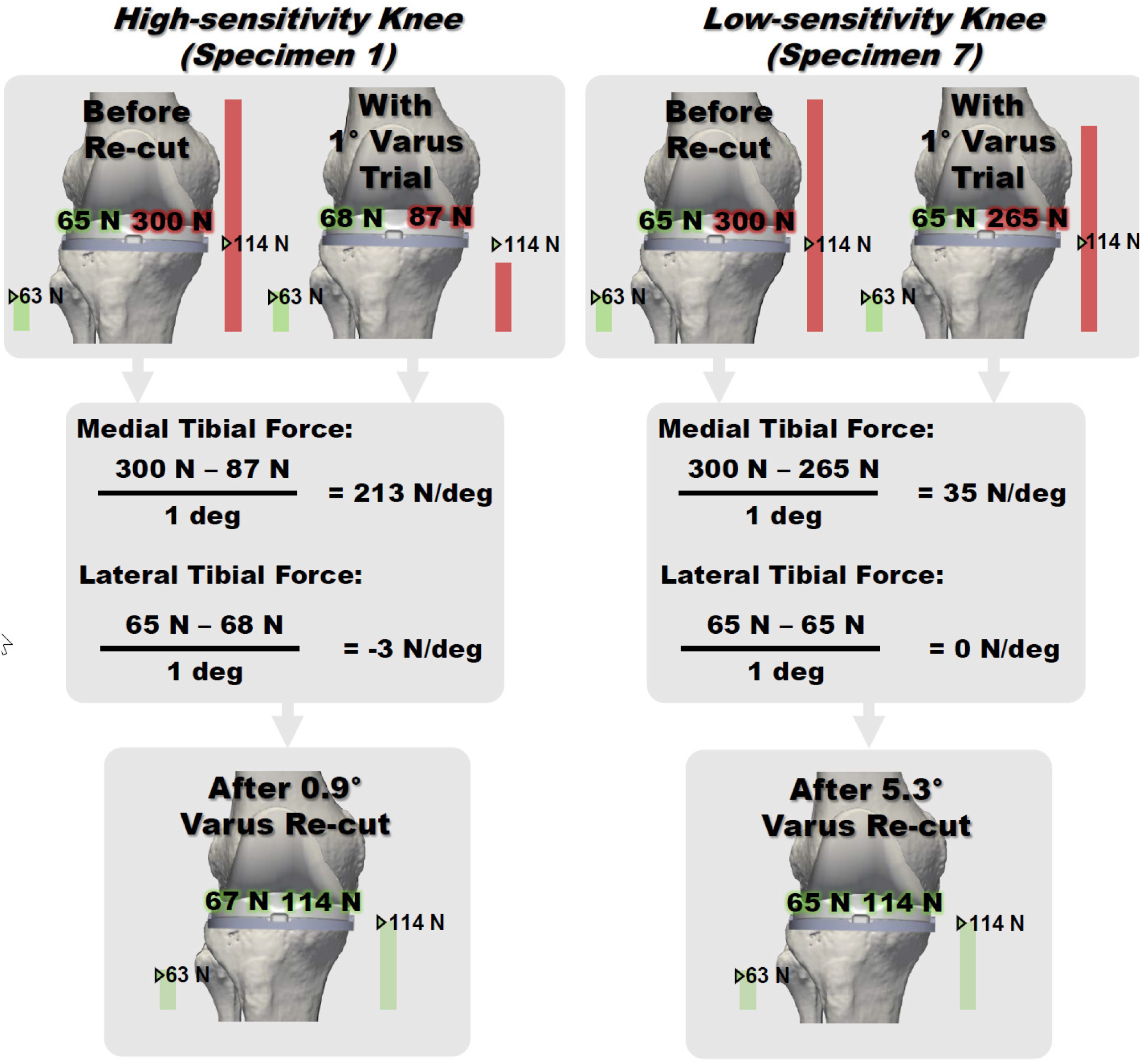
Schematic shows representative examples of how patient-specific sensitivities can be determined using a small trial re-cut (e.g., 1° varus) and then how the correct re-cut for that patient can be determined based on the patient-specific sensitivity. Because the target tibial forces for a specific patient are unknown, the mean tibial forces at 0° flexion in the native knee (medial force = 114 N and lateral force = 63 N) ^13^ are used as the target for this example.

Three methodological issues should be acknowledged because of their potential to affect the results. One methodologic issue concerns the alignment of the tibial component in the reference condition. Tibial component malalignments in this reference condition relative to the native joint line might change the initial tension in the soft tissue restraints. A pair of recent studies showed that (1) the tibial forces after KA TKA are comparable to those of the native knee^20^ and (2) the laxities and neutral positions were largely not different from those of the native knee^22^. Thus, if present, then V-V malalignments of the reference tibial components were likely small as described previously in the Methods section^2^. A second methodologic issue was that this study was performed in knees without OA. Thus, the sensitivities might differ in knees with end-stage OA. Additional changes to the mechanical properties of the medial and lateral soft tissue restraints that likely occur in some OA patients would likely increase the variability in the sensitivities. However, while the exact sensitivities might change in OA knees, the conclusions drawn in the present study that tibial forces are better than laxities for detecting overstuffing should still apply to OA patients. Further, using the simple patient-specific approach described previously would enable the patient-specific sensitivity to be determined regardless of the condition of the soft tissue restraints. The third methodological issue is that only nine knee specimens were included. While this number was sufficient to detect strong relationships between V-V malalignment and changes in tibial forces and laxities, this group likely does not capture all the knee-to-knee variability in the population. To best account for this issue, we computed tolerance intervals to represent the predicted range because tolerance intervals take into account the sample size when predicting the range of the population.

In conclusion, measuring changes in tibial forces is more useful than measuring changes in laxities for identifying and correcting V-V malalignments of the tibial component that overstuff a compartment. Because of the wide variability in sensitivities between knees, a patient-specific procedure is needed to adjust the tibial resection when high tibial forces are registered due to overstuffing of a compartment. Using the simple patient-specific procedure proposed in this study, surgeons can determine patient-specific sensitivities of tibial forces to V-V malalignments of the tibial component and use these patient-specific sensitivities to determine the re-cut necessary to correct overstuffing caused by V-V malalignment of the tibial component. These patient-specific re-cuts should reduce the risk of improperly tensioned soft tissue restraints and unnecessary ligament releases.

## Data Availability

Please contact the corresponding author with requests for data.

## Acknowledgements

The authors acknowledge the support of the National Science Foundation (Award No. CBET-067527) and Zimmer Biomet (Award No. CW88095). Additionally, the authors wish to thank individuals who donate their bodies and tissues for the advancement of education and research.

One author, MLH, received research funding from Zimmer Biomet for this study and receives research support from Medacta that is not related to this study. One author, SMH, receives IP royalties from and is a paid consultant and presenter/speaker for Biomet Sports Medicine and Medacta, and receives research support from Medacta, which are all unrelated to this study.

**Supplement 1: Illustrative example of difficulty with using mean sensitivities to guide balancing KA TKA**

To appreciate the difficulty with using the mean sensitivities to guide corrections in all knees, consider an illustrative case in which the surgeon measures a medial tibial force of 300 N and a lateral tibial force of 65 N in extension. Because the ideal tibial forces for a particular patient are unknown, it is reasonable to strive to achieve tibial forces comparable to the average in the native knee^13^, which are similar to those measured in calipered kinematically aligned TKA *in vitro*^20^. Using the mean tibial forces of the native knee as a target (114 N in the medial compartment and 63 N in the lateral compartment^13^), the medial force is 186 N too high, and the lateral force is 2 N too high, which indicates that there is a valgus malalignment of the tibial component (Figure 2, Table 1). Thus, a varus re-cut should correct the valgus malalignment and reduce the medial tibial force with minimal changes to the lateral tibial force (Figure 2, Table 1). However, the appropriate size of the varus re-cut remains to be determined.

To understand the importance of considering knee-to-knee variability in the sensitivities of the tibial forces, first assume that this patient has sensitivities equal to the highest determined in the present study (213 N/deg valgus malalignment, specimen 1, Table 3). If the surgeon used the average sensitivity to determine the appropriate re-cut, then a varus re-cut of 1.6° would be performed (186 N / (118 N/deg) = 1.6°). However, in this high-sensitivity knee, a 1.6° varus re-cut would be much too large because the actual decrease in the medial force would be 336 N. Because a decrease of 336 N would be greater than the measured medial force, this re-cut would likely slacken the soft tissue restraints in the medial compartment, which might cause excessive laxity post-operatively. Next, assume that this patient has sensitivities equal to the lowest in the present study (35 N/deg valgus malalignment, specimen 7, Table 3). In this low-sensitivity knee, the 1.6° varus re-cut would be too small because the actual decrease in medial tibial force would only be 56 N. In both these patients, additional surgical steps would be necessary to achieve tibial forces comparable to those of the native knee, which is undesirable due to increasing time and complexity of the procedure.

